# Experiences of children’s self-wetting (including incontinence) in Cox’s Bazar’s Rohingya refugee camps, Bangladesh

**DOI:** 10.1101/2023.08.21.23294365

**Authors:** Mahbub-Ul Alam, Sudipta Das Gupta, Claire Rosato-Scott, Dewan Muhammad Shoaib, Asmaul Husna Ritu, Rifat Nowshin, Md Assaduzzaman Rahat, Nowshad Akram, Joanne Rose, Barbara E. Evans, Dani J. Barrington

## Abstract

Self-wetting, including incontinence, affects people of all ages, ethnicities, and cultural backgrounds, and can have a significant negative impact on quality of life. We thus explored the attitudes towards self-wetting and experiences of children (ages five to 11), their caregivers, and humanitarian experts in the Rohingya refugee camps in Cox’s Bazar, Bangladesh.

We purposively selected participants from two camps where our partner organisation, World Vision Bangladesh - Cox’s Bazar, works. We conducted Key Informant Interviews (KIIs) with community members and camp officials, Story Book (SB) sessions with Rohingya children and in-depth Interviews (IDIs) with caregivers of children who participated in the SB sessions, as well as surveying the communal toilets used by children of the caregivers.

Self-wetting was commonly seen among the children. Due to self-wetting, children were likely to feel embarrassed, upset and uncomfortable, and frightened to use the toilet at night; many also indicated that they would be punished by their caregivers for self-wetting. Key informants indicated that caregivers have difficulty handling children’s self-wetting because they have a limited amount of clothing, pillows, and blankets, and difficulty cleaning these items. In the sanitation survey it was evident that the toilets are not appropriate and/or accessible for children.

Children in the Rohingya camps studied self-wet due to both urinary incontinence (when unable to reach a toilet in time) and because the sanitation facilities offered are inappropriate. They are teased by their peers and punished by their caregivers. The lives of children who self-wet in these camps could be improved by increasing awareness on self-wetting to decrease stigma and ease the concerns of caregivers, as well as increasing the number of toilets, ensuring they are well-lit, providing child-friendly toilets and cubicles, fixing the roads/paths that lead to sanitation to facilities and increasing the provision of relevant continence management materials.

## Introduction

The medical condition of urinary incontinence (UI) is defined as the involuntary loss of urine [1], and it is a complex global health issue that has a negative impact on people’s security, dignity, rights and general quality of life [2, 3]. People who experience UI, and their caregivers, face challenges in their everyday lives, with individuals reporting that the intensity and complexity of the experience changes on a daily basis. Incontinence can also result in social and economic marginalization, debilitation, and psychosocial problems due to the associated stigma [4]. The stigmatization of UI prohibits individuals from sharing their difficulties with others, and because of that, they often separate themselves from society, community, and family [4].

Children aged between five and 11 years occasionally wet themselves as a result of the medical condition of UI; when this happens during sleep, it is referred to as enuresis [5]. Sometimes children of this age wet themselves because they do not want to, or cannot, access a toileting facility in time; not due to a physiological inability to stop urine leakage. This is known as social incontinence. The term ‘self-wetting’ can be used when a child wets themselves, but the cause is unknown. Regardless of whether self-wetting is physiological or due to a lack of appropriate facilities, the shame and humiliation linked with it can have an impact on relationships and involvement in social events, increasing the likelihood of psychological difficulties in childhood. Increased domestic violence towards children who experience enuresis has been observed [6] and children who self-wet can also suffer from skin rashes and Urinary Tract Infections (UTIs) [7]. The consequences of self-wetting mean that children who self-wet usually try to conceal the condition, although daytime UI is difficult to conceal, particularly where continence aids such as absorbent pads are not available [8]. Children who do not achieve continence ‘on time’ (according to culturally specific expectations of achieving toilet training) can suffer long-term psychological issues [8].

Very little work has investigated the prevalence and experiences of self-wetting in low resource settings [3, 9], but the available data suggest that it is common around the world, regardless of culture, age and ethnicity [10–13], and that experiences in these settings are poor [14–17]. To the best of our knowledge, the prevalence of childhood self-wetting, including UI, has not been successfully measured in low resource settings, with a recent study that attempted to do so noting that the stigma associated with self-wetting makes it a challenging task to identify children who experience it [18].

In humanitarian settings, self-wetting by all age groups often goes unnoticed [17]. Yet the incidence of UI may be higher in humanitarian settings because of situation-induced trauma, anxiety and physical harm [19, 20], and self-wetting generally due to the decreased lack of access to appropriate toileting facilities. For example, International Rescue Committee (IRC), Médecins Sans Frontières, International Federation of Red Cross and Red Crescent Societies (IFRC) and the United Nations High Commissioner for Refugees have observed an increase in bed-wetting among children in Syria, Lebanon, Iraq, Greece and Honduras since the onset of fighting and/or their displacement [17]. Jurkovic, et al. (2019) found that refugee or displaced children have an increased risk of incontinence, which can cause further trauma in itself. Families in emergency situations face additional challenges in managing their children’s self-wetting due to a lack of resources, including water and soap [17]. Self-wetting limits the accessibility of essential services (food, water, and health care) and the opportunity to participate effectively in decision-making processes, leading to further social marginalization and vulnerability [3, 9, 17].

There is a lack of knowledge on the challenges and obstacles that children who self-wet and their caregivers encounter in emergency settings, and how support to manage the condition can be effectively provided in the planning, implementation and assessment of humanitarian programming. Our study took a phenomenological approach using a Story Book (SB) methodology (development of the methodology is detailed in [21]) in-depth interviews and a sanitation survey based on the CHILD-SAN approach [22] to understand the barriers to inclusion and well-being that those living with self-wetting, particularly children aged five to 11 and their caregivers, face in the Rohingya refugee camps of Cox’s Bazar, and how more holistic, effective and inclusive WASH and protection programming can be developed to support those who self-wet and their families.

## Methods

We report this study in line with the consolidated criteria for reporting qualitative research [23] (checklist included as Supplementary Information 1).

### Study Setting

This study was conducted in the Rohingya refugee camps of Cox’s Bazar, Bangladesh, the world’s largest refugee settlement, inhabited by over 950,000 refugees/Forcibly Displaced Myanmar Nationals. Children comprise more than half of the population, with those aged five to 11 years comprising 11% [24]. A concurrent study using the same methodology was conducted in refugee settlements in the Adjumani District of Uganda; results of that study are forthcoming.

### Study Design

We used a phenomenological approach to understand lived experiences of children’s self-wetting in this context. We undertook SB sessions with Rohingya children aged five to eleven years old, in-depth interviews (IDIs) with their caregivers, key informant interviews (KIIs) with community members (camp leaders, religious leaders and traditional healers) and camp officials (teachers, community health workers, child protection officers and WASH specialists) and surveyed sanitation facilities used by the children and their family members for their appropriateness to children of this age group [22]. We then triangulated across these methods to understand lived experiences and possible ways to improve them.

The SB methodology was developed by the research team to hear from children aged five to 11 years old about how an imaginary ‘hero’ character, approximately their age and living in one of the Cox’s Bazar camps, might experience self-wetting. Children were asked to express their understanding, experiences and feelings of facing self-wetting issues through their drawings and discussions. The methodology was developed using a participatory process with local contextualization and is discussed and evaluated elsewhere [21]. All of the data collection tools were developed by the research team and Advisory Committee members (specialists on research with children, incontinence and emergency contexts) and then reviewed and finalized in coordination with the local research team in Bangladesh. Questions were translated into Bangla and then verbally adapted into the Rohingya language while conducting activities. All of the tools used are available in English and Bangla via the Open Science Framework [25].

### Participant Recruitment

Two research sites (Site 1 and Site 2) were selected based on accessibility, availability of the children and caregivers and pre-existing relationships between the local research partner (World Vision Bangladesh - Cox’s Bazar (WVB-CXB)) and the Rohingya community. Children known to self-wet were not purposively recruited, as the local research team believed this may increase stigma and potentially protection risks. Instead, interested children already known to WVB-CXB (and who may or may not self-wet) were invited to participate during a community visit, with the understanding that even children who do not self-wet may have insights into how those who do are treated (including by themselves) within their community. We conducted eight SB sessions with 48 children, one at each research site with girls aged five to seven years old, boys aged five to seven years old, girls aged eight to 11 years old, and boys aged eight to 11 years old. To understand the caregivers’ experiences and understanding of self-wetting by children, we invited (face to face) 12 caregivers of children the SB moderator identified as likely experiencing self-wetting and a further 12 who likely were not, and interviewed them. These 24 caregivers were purposively selected from the eight SB sessions to represent children of different age groups and gender. To further understand the challenges of self-wetting in children and discuss possible solutions to issues raised during the IDIs and SB sessions, we conducted KIIs with individuals who are engaged in the day-to-day care of children living with self-wetting and/or directly involved in addressing issues of self-wetting based on their positions or roles. We invited three teachers, two Community Health Workers (CHWs), three camp leaders, one religious leader, two traditional healers, two child protection officers, and two WASH specialists face to face, by emails or by phone calls, and interviewed them as key informants (Table 1).

**Table 1:** Overview of Participant Groups by data collection method.

| **Participant Group** | **Data Collection Method** | **Site** | **Number of Activities** | **Male Participants** | **Female Participants** | **Total Participants** |
| --- | --- | --- | --- | --- | --- | --- |
| Boys aged five to seven years old | Story Book session | 1 | 1 | 6 | 0 | 6 |
|  |  | 2 | 1 | 6 | 0 | 6 |
| Girls aged five to seven years old | Story Book session | 1 | 1 | 0 | 6 | 6 |
|  |  | 2 | 1 | 0 | 6 | 6 |
| Boys aged eight to eleven years old | Story Book session | 1 | 1 | 6 | 0 | 6 |
|  |  | 2 | 1 | 6 | 0 | 6 |
| Girls aged eight to eleven years old | Story Book session | 1 | 1 | 0 | 6 | 6 |
|  |  | 2 | 1 | 0 | 6 | 6 |
| Caregivers | IDI | 1 | 12 | 0 | 12 | 12 |
|  |  | 2 | 12 | 0 | 12 | 12 |
| Teachers | KII | 2 | 2 | 0 | 2 | 2 |
|  |  | 1 | 1 | 0 | 1 | 1 |
| Community Health Workers (CHW) | KII | 1 | 2 | 2 | 0 | 2 |
| Camp Leaders (Majhi) | KII | 1 | 1 | 1 | 0 | 1 |
|  |  | 2 | 2 | 2 | 0 | 2 |
| Religious Leaders | KII | 2 | 1 | 1 | 0 | 1 |
| Traditional Leaders | KII | 1 | 1 | 1 | 0 | 1 |
|  |  | 2 | 1 | 1 | 0 | 1 |
| Camp Child Protection Officials (CPO) | KII | 2 | 2 | 1 | 1 | 2 |
| WASH Specialists | KII | 1 | 1 | 1 | 0 | 1 |
|  |  | 2 | 1 | 1 | 0 | 1 |

### Research Training

DJB (PhD, female, Lecturer in Global Health) and CRS (MSc, female, PhD candidate) led the project across both Uganda and Bangladesh, and recruited MUA (MPH and MSS, male, associate scientist) to oversee research training and data collection in Bangladesh due to his experience as a qualitative researcher in the field of WASH. MUA and SDG (MSS, male, research officer) provided three days of training to the six data collectors (three male and three female WVB-CXB staff who have experience conducting participatory discussions with children) on the background and purpose of the research and the principles of qualitative data collection, including the data collection techniques of KIIs, IDIs, the SB methodology and the sanitation survey, and ethical considerations.

### Data Collection

The research team recruited participants and collected data from October 10 to October 25, 2021. SDG and the WVB-CXB staff collected data in the local language/s. The SB sessions were facilitated by a data collector of the same gender as the children and took place in school classrooms; caregivers were sometimes visibly present but not within earshot of the children’s discussions. The SB sessions took between 50 and 155 minutes. Caregivers were interviewed in their households, as per their preference. The KIIs took place at the individuals’ work/living place, including formal workplaces, households, schools, and mosques, based on their preference. The IDIs and KIIs were between 30 and 60 minutes long. All SB sessions, IDIs and KIIs were audio recorded, and photographs of children’s drawings taken during SB sessions. We also surveyed the sanitation facilities available to the children of the caregivers we interviewed. These children and their family members used these sanitation facilities on a daily basis. We visited the available communal sanitation facilities and assessed the status of accessibility issues, toilet walls, toilet roof, toilet door, door handle, door lock, handwashing water container, soap, hygiene promotion and available facilities within the toilets, using an observational checklist [25] (available at Barrington, 2023).

### Ethical Considerations

This is a very sensitive topic, and from the outset we had to consider whether it was ethical to conduct such research at all in an emergency setting, particularly the Story Book sessions which involved children. For an in-depth discussion on our considerations and eventual decision to conduct the work see Supporting Information 1 of Rosato-Scott et al. [21]. Approval to conduct the project was granted by the Research Ethics Committee, Faculty of Engineering, University of Leeds, United Kingdom (Reference MEEC 19-020). Approval to conduct the research in Cox’s Bazar was granted by the Institutional Review Board of the Institute of Health Economics (University of Dhaka, Bangladesh), with authority to access the refugee camps granted by the Office of the Refugee Relief and Repatriation Commissioner (RRRC). The research team explained to all participants that they wanted to better understand how children aged five to 11 years old experience self-wetting, and whether there are ways to improve these experiences (participant information sheets are available from [25]). We obtained verbal assent from the children for the SB sessions, and written consent from their caregiver. For the KIIs and caregiver IDIs, we obtained written or verbal (where a second data collector acted as a witness and signed the form) informed consent.

### Data Analysis

All activities were transcribed and translated verbatim into English; participants were not asked to comment on or correct them. Data collectors did not take formal fieldnotes, but SDG debriefed the local research team daily and discussed potential probes for SB sessions and interviews that could be used to identify and explore emerging themes. MUA and SDG undertook initial data analysis during the data collection stage.

After the completion of data collection, following a deductive approach, MUA, SDG, DJB and CRS initially developed a coding framework based on the research objectives. Inductive codes were developed using constant comparative analysis as the work progressed. SDG, AHR, RN, and MAR coded all transcripts using NVivo 12 (QSR International), dividing them among themselves equally, and coding of all transcripts was then checked by DMS and MUA. Data were triangulated between the data collection methods. Although it was originally planned that initial findings would be discussed with participants to incorporate their feedback into a final round of analysis, restrictions related to COVID-19 and the funding period meant this was not possible.

## Results

The SB sessions, IDIs, KIIs and sanitation survey provided insight into three major facets of self-wetting by children in humanitarian contexts: the perceived causes of self-wetting, experiences of children and caregivers when managing the condition, and suggestions for reducing the incidence of self-wetting (See S2 Table for the full codebook).

### Perceived causes of self-wetting in children

#### Self-wetting as a ‘disease’ or a normal part of life

Participants indicated that the Rohingya community usually refer to self-wetting as “*Korai*”, and it is common among children aged five to 11 years. Several participants, including caregivers (6 of 24), CHWs (2 of 2), majhis (local leaders among Rohingyas, 2 of 2) and a religious leader (1 of 1), consider self-wetting and the medical condition of UI a ‘disease’. As a group, they consider self-wetting a disease because they believe it needs treatment to be cured. Those caregivers who consider it a disease do so because children urinate in their sleep on a daily basis and continue to do so even after consulting with doctors or Hakeem. A CHW stated that “We call it [self-wetting] a disease because [we can see] when people or children sleep during the daytime and dream, they sometimes defecate/urinate in their sleep. Who [children] cannot control their urge to defecate, whether the toilet is far away or near, they urinate in bed or on clothes when they cannot control it. Others lose control on the way [to the toilets] because the bathroom is far away, unable to hold it all the way.” (CHW 2) and one caregiver “If the child is sick, feels troubled, and urinates in bed, won’t the mother feel troubled? He [the boy child] is urinating in bed because he has a disease.” (Caregiver of Child 3 from SB Session 1).

A few caregivers (5 of 24), but several KIs (10 of 15), consider self-wetting a normal phenomenon which resolves with age. They feel that as children become older, they gain bladder control; children self-wet when they are still too young to sufficiently control their urine, not because they are ill. OneCHW and both CPOs believe children usually urinate in bed while deep asleep because they are dreaming of urinating in toilets. Children suggested that major reasons their heroes wet themselves at night are excessive intake of water before going to sleep (8 of 8 sessions), and an inability to control the urge to urinate while dreaming (4 of 8 sessions). None of the health care service providers, or the CPOs, are informed about the self-wetting issues of children as the issue is not reported to them by the community.

#### Available toilets are inappropriate

The most common reason participants mentioned for children self-wetting was that children consider the toilets available to them are inappropriate for their needs. Some of the caregivers mentioned that children are afraid to use the toilet at night because the toilets are dark (6 of 24), far away (12 of 24), dirty (5 of 24), and the roads to toilets are in a poor condition (10 of 24). A major reason children gave for their hero self-wetting was distance from the toilet (5 of 8 sessions). From the sanitation survey, it was evident that the communal toilets available to the children participants are not all appropriate. We only deemed 4 of the 24 toilets ‘child friendly’. Nine of the toilets are missing door handles and 20 do not have locks. Caregivers reported that the toilets are also frequently broken and the doors need to be closed using wire, which discourages children. As a result of the many inappropriate aspects of the sanitation available to them, children often urinate or defecate in their bed or on themselves or their mat. The sanitation survey also revealed that the majority of the toilets are far from households, taking five to ten minutes to get to, and require crossing a sloppy, muddy road. There is no signage to ten of the toilets and difficult-to-see signage for the other 14. Most of the paths (23 of 24) to the toilets do not have any lighting.

In all the SB sessions the children indicated that their hero did not feel comfortable and safe using the toilet. One girl aged eight to 11 years explained that their hero wet the bed because “The latrine is far away; it is scary to go there. That is why it [hero] is late to go there.” (SBSession 6). Caregivers stated that “Child feels scared to go to the bathroom at night; there are no lights. The road to the toilet is not good either; the road is dark.” (Caregiver of Child 4 from SB Session 6) and “The path to the toilet is not friendly. Now where it is, it is pretty low [in position/placement]. Now which [the new toilets] are going to be newly formed, those should be formed in plain land. Then there will be no more difficulties, and the kids will not fall anymore. But the kids are suffering there now”. (Caregiver of Child 4 from SB Session 2).

Caregivers (13 of 24) observed that even when children can reach the toilet they often do not use the toilet because they are generally not child friendly. Children aged five to 11 years cannot sit properly on the toilet seat and face problems in using the water for anal cleansing and flushing. One WASH specialist explained that “latrine’s size, pan’s size, people’s structures, 5 feet, 6 feet, like this. But children’s body structure is small. If their pan is 34 inches, 8 inches on this side, they cannot sit with two feet on two sides. It becomes difficult. If we make a child sit on an adult’s pan, then s/he will not be able to do it properly. If his legs are spread too wide, he won’t feel that pressure.” (WASH Specialist 1).

Each block in the Rohingya camps has one toilet that is shared by four or five households, resulting in lengthy wait periods that further deter children from using them. Caregivers (9 of 24) mentioned that sometimes, children have to wait for their turn to access the toilet while adults are using it, meanwhile urinating while unable to hold it. Some of the service providers (4 of 10) confirmed that a long waiting time to get to the toilet is one of the significant causes of self-wetting by children. In addition, because of the volume of users, the available toilets are unclean and children do not want to use them. As two caregivers stated “This latrine becomes very unclean. There are a lot of people here. There are 8/10 people or 10/12 people in each family. For this, the latrine becomes very unclean. We have to clean it; it smells bad if we don’t clean it. That’s why we have to clean it. Children don’t want to use it if it’s unclean.” (Caregiver of Child 4 from SB Session 6) and “I thought it would be nice to have another latrine for the children. We [the adults] use this latrine, [along with] 3/4 families, together. they [children] don’t want to go to unclean latrines.” (Caregiver of Child 3 from SB Session 5).

### Experiences of self-wetting in children

#### Children are likely distressed when they self-wet

In all of the SB sessions children indicated that their heroes felt uncomfortable, angry, scared, tense, and embarrassed after self-wetting. It seemed that through their drawings the children were expressing their own feelings about self-wetting (regardless of whether it is something they experience or not). The children often stated that their hero cried and felt distressed after an incident. One of the main reasons their heroes were scared is that they believed their (heroes’) mother would scold them because she was upset, furious, sad and/or embarrassed (7 out of 8 SB sessions). For example:

> *“Facilitator: How does your heroine feel when her clothes get wet, dear?*

> *Respondent 1: She feels troubled. And also feels scared that her clothes were wet, her mother would scold her……*

> *Respondent 2: She urinates in bed; that’s why she feels terrible and ashamed…..*

> *Respondent 3: She feels sad.”*

> *(SB Session 6)*

The children particularly illustrated this when discussing or drawing what would happen when their heroes got out of bed in the morning and had wet themselves. They indicated that caregivers sometimes slap the children or beat them with whatever they can find nearby, such as a broom or stick (discussed in 6 of 8 SB sessions), or by grabbing their hair (5 of 8 SB sessions) (Figures 1 and 2). Some children chose to disclose that this had happened to them.

> *“Facilitator: Suppose one day, the heroine accidentally urinated and wet the bed. How will she feel?*

> *Respondent 3: She will be ashamed*.

> *Respondent 1: She will be scared*.

> *Facilitator: You said that the heroine does not feel good; she cries, she feels scared and sad. Why does she feel like this?*

> *Respondent 1: Her mother will beat her.”*

> *(SB Session 3)*

**Figure 1:**
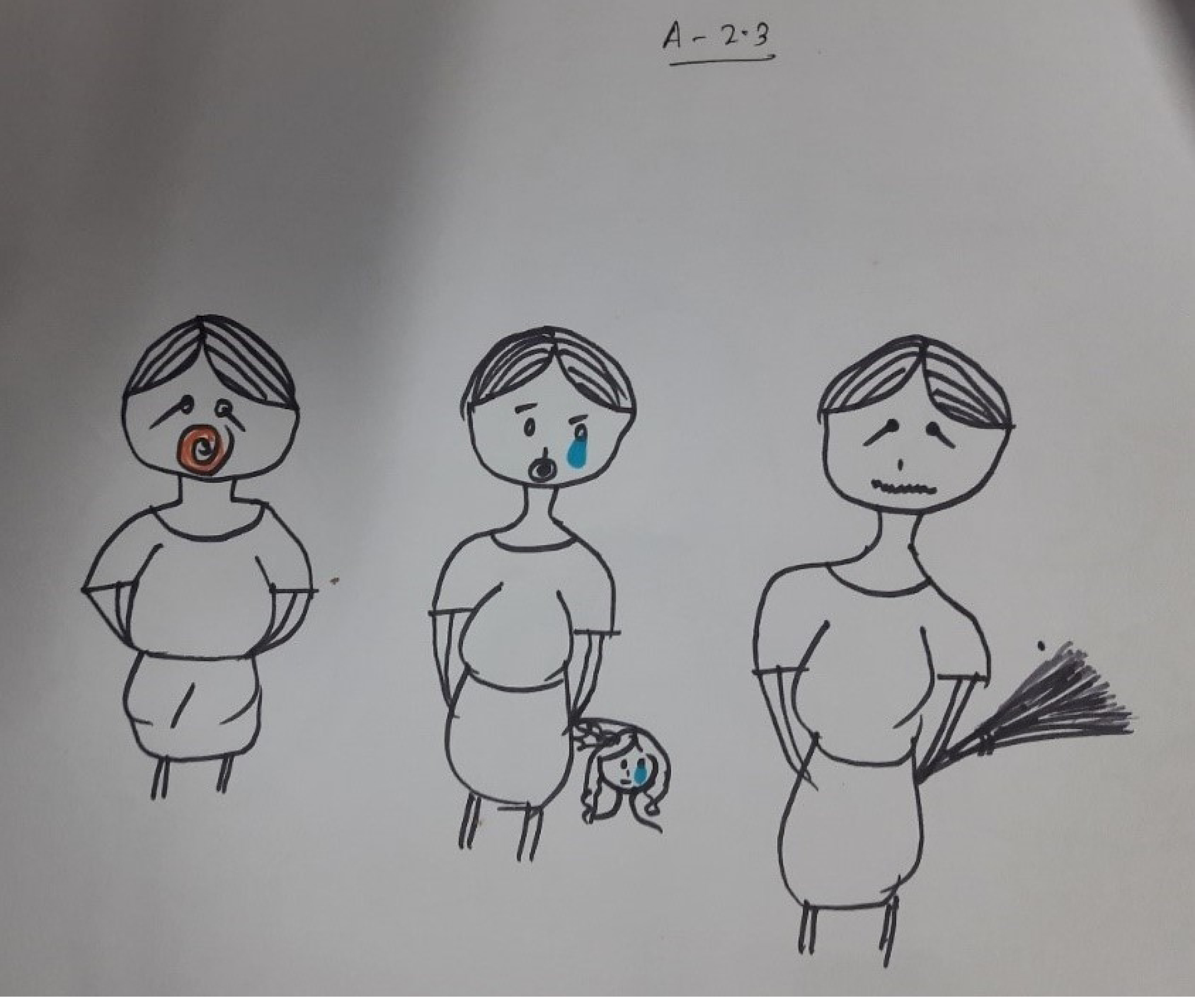
Drawing of the hero’s mother scolding the child, crying and beating the child with a broom, as well as the hero crying. Story Book Session 6, Girls 8 to 11 Years Old, Site 01 (Authors’ own photograph)

**Figure 2:**
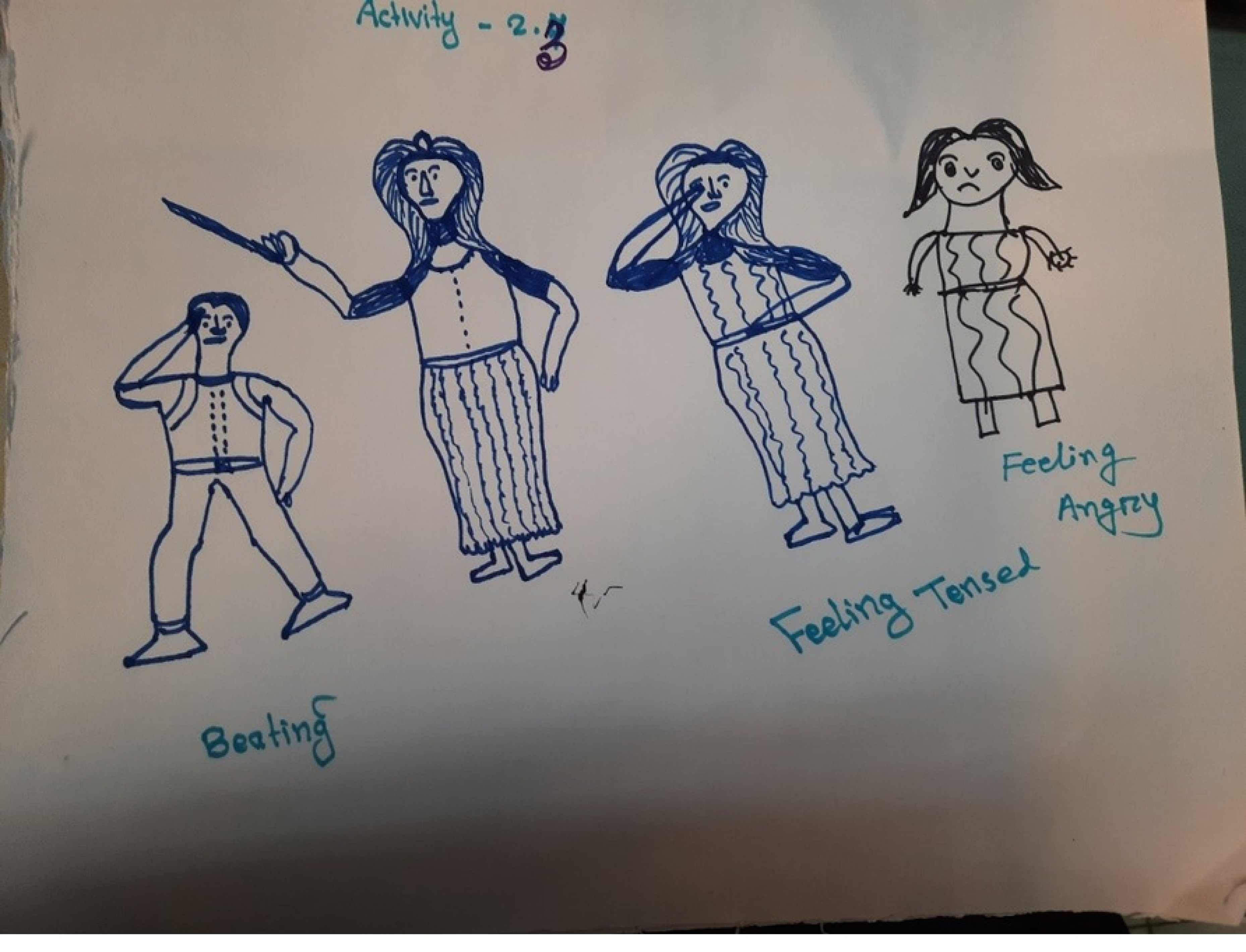
Drawing of a mother hitting the hero for self-wetting, being tense and being angry. Story Book Session 1, Boys five to eight years old, Site 2 (English words added by data collector from description provided by illustrator) (Authors’ own photograph)

In half of the SB sessions children mentioned that teachers and classmates mock and ridicule children who self-wet during school. Children explained that sometimes teachers are compassionate and advise students to go home and change their clothes after urinating in them, but others frequently become angry and beat the students for making their clothes wet at school (4 of 8 SB sessions). They either remove those children from school or call their fathers.

> *“Facilitator: She (heroine) urinated after going to school. What will everyone in the school do? What will the teacher do?*

> *Respondent 1: They will make her feel ashamed.”*

> *(SB Session 6)*

> *“Respondent 1: The teacher beat him (hero)*.

> *Facilitator: The teacher beat him. Didn’t the teacher do anything else?*

> *Respondent 3: The teacher drove him away. The teacher kicked him out of school.”*

> *(SB Session 7)*

#### It is challenging for some caregivers to manage their children’s self-wetting

Dealing with their children’s self-wetting issues makes some caregivers anxious (5 of 24), uncomfortable (4 of 24), and ashamed (2 of 24). When children urinate or defecate in their clothes or bedding, caregivers (particularly mothers) are normally the ones who wash the clothes and linen, which annoys them and sometimes results in punishment of the children; “My son is urinating and defecating; I have trouble washing their clothes. I beat the children and changed their clothes. I feel sad about that.” (Caregiver of Child 5 from SB Session 6). In the community, children who experience self-wetting and their parents feel ashamed as they face humiliation from the community because of their children’s open urination and defecation.

Some caregivers (3 of 24) mentioned the burden of managing their children’s self-wetting. They have to fetch water from a distance to clean the clothes and the child because there is no well near their homes. One CPO, one CHW and one religious leader also mentioned that the lack of water availability nearby, due to the humanitarian context, likely contributed to this stress. Several service providers (4 out of 10) mentioned that caregivers face challenges in managing children’s self-wetting as they have a limited amount of clothes, pillows, and blankets, and if anyone urinates or defecates on the beds, they face problems using and cleaning these.

#### Caregivers of children who self-wet seek help

To treat self-wetting, caregivers usually do or would seek help, either from religious leaders (Moulovi) (19 of 24) or doctors (12 out of 24). Many caregivers (9 of 24) prioritize traditional/spiritual healers over doctors. The spiritual healers or ‘Moulovis’ recite the Quran, provide amulets, sacred water, or oil to treat this incontinence issue. As one caregiver explained “Parents have two ideas [when observing incontinence]. I think it would be better to go to the doctor first. [However,] another idea is that [going to the religious leaders]. They believe it happens because of dreams or supernatural jinn ghosts. “ (Caregiver of Child 3 from SB Session 1).

### Suggestions for reducing self-wetting

#### Participants believe that self-wetting can be prevented

A few of the caregivers (4 of 24) felt that self-wetting could be prevented if there was better access to doctor’s treatments in the camps. The children suggested that to prevent self-wetting their heroes could drink less water (4 of 8 SB sessions) and/or urinate before going to sleep at night (4 of 8 SB sessions).

The majority of other recommendations of participants were around improving WASH facilities. Caregivers and key informants suggested that to decrease self-wetting more toilets should be built in general (4 out of 24 caregivers, 9 of 18 KIs), and these should be well built (6 of 24 caregivers, 7 of 18 KIs), nearer to households (6 of 24 caregivers, 9 or 18 KIs), have water available and the roads or trails to them improved (3 of 24 caregivers, 8 of 18 KIs), with availability of lights at night (6 of 24 caregivers, 8 of 18 KIs). A CHW (1 of 2) and CPO (1 of 3) suggested providing torches for children to use at night when walking to the communal toilets.

Children in half of the SB sessions felt that more toilets needed to be built closer to or in homes. Some caregivers (4 of 24) and two Majhi (2 of 3) suggested providing a small area at the household for the children to urinate and defecate, “Even if you cannot arrange a bathroom, there should be a small place for the children (at their house) to urinate and defecate. For example, if the older adults cannot go out, their toilet chairs are arranged.” (KII_School_Teacher_03).

Both WASH specialists suggested that smaller commodes (e.g., smaller pan) should be provided for children so that they can use toilets more easily; “small pans need to be done for children so that s/he can sit comfortably.” (Wash Specialist 1)

## Discussion

Self-wetting can lead to negative physical [7], social [2, 3] and mental [6, 8] health impacts, particularly for children. Our study revealed that the uncomfortableness and embarrassment the children face because of wetting their clothes and bedding through accidental urination and defecation lead them to be angry, scared and tense. Children are often afraid of their mother’s rebuking and being beaten because of wet clothes. A study conducted in Brazil showed that 89% of the participating children experiencing continence issues were abused verbally while 49% were punished physically, and in 88% of the cases the primary abuser was their mother [26]. Children also experience reduced sleep quality because of the anxiety and fear of possibility that they might leak during sleep [27].

The frequent urination or defecation of children in bed at night also contributes to feelings of the caregivers being upset, furious, sad, and embarrassed. The caregivers face humiliation from the neighbors and community members because of their children’s self-wetting. This feeling can lead them to punish the children, as has been observed elsewhere [9].

Findings from this study highlight the difficulties children who self-wet and their caregivers face in their personal, social, and academic life as a result of the concealed nature and stigma surrounding these issues. The abuse associated with incontinence that the children face in the form of mockery and ridicule from teachers and classmates impacts them. Stigma results in embarrassment and shame, which discourages children from participating in program, educational and social activities [28, 29].

The children also face physical violence from some teachers because they wet their clothes at school, which makes some teachers angry, whilst other teachers advise children to change their clothes or go home. It has been demonstrated elsewhere that children are often at risk of abuse from teachers because of self-wetting [6].

Through this research we have developed a causal model to explain how these negative wellbeing impacts occur through children’s, caregivers and key informants’ experiences of self-wetting in the Rohingya refugee camps (Figure 3). There are three ‘pain points’ in the model where we believe WASH and protection practitioners can remove or reduce factors which contribute to poor well-being, thus improving the experiences of children and their caregivers. These are improving WASH facilities, providing continence management supplies and increasing knowledge of self-wetting whilst reducing stigma. Medical intervention may also be required in some instances, but this should be delivered by health specialists, not WASH and protection practitioners.

**Figure 3:**
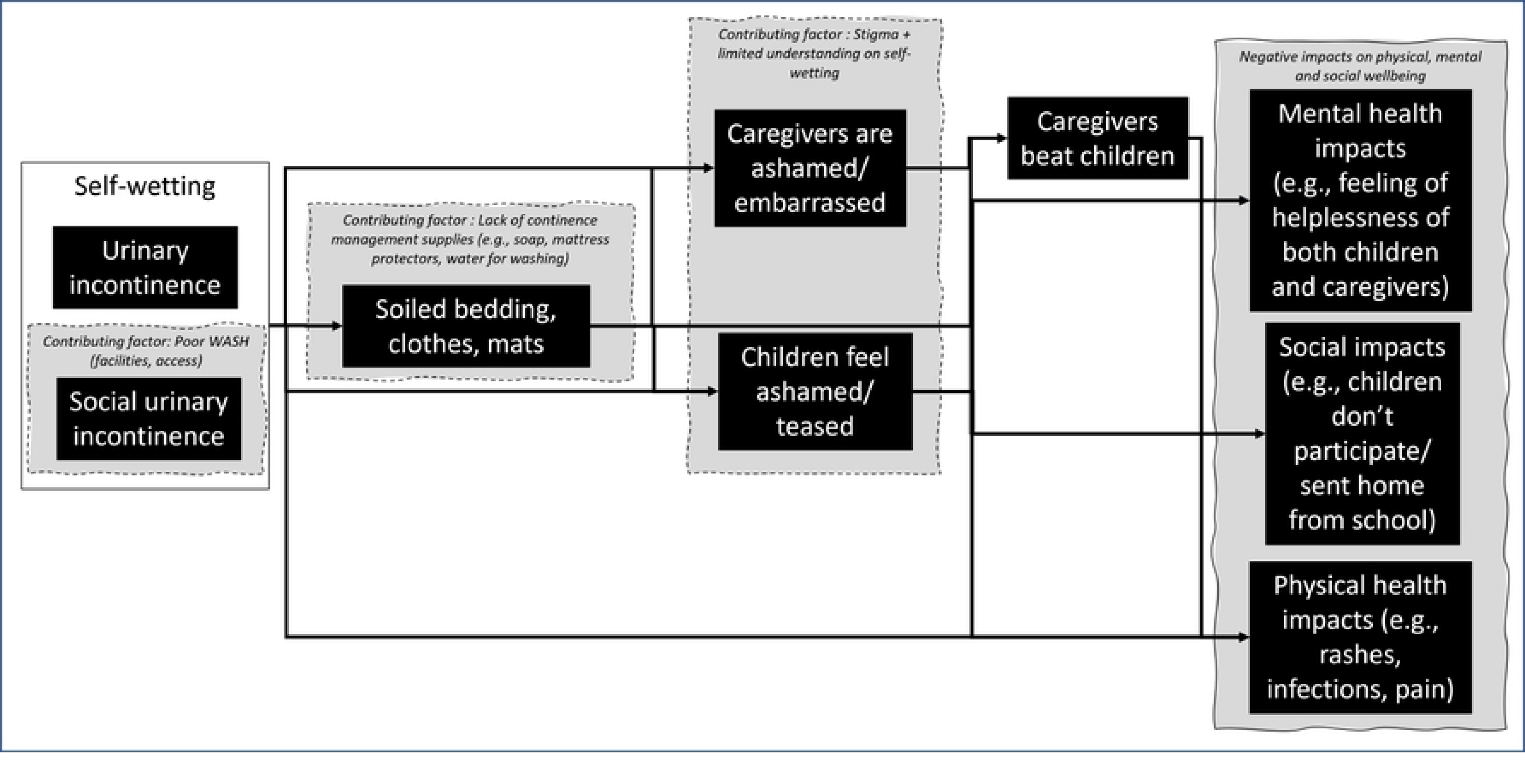
Model of self-wetting experiences of children aged 5-11 years in Rohingya refugee camps.

### Water, Sanitation and Hygiene facilities

The accessibility issues at and leading to the communal ablution blocks contribute to children experiencing self-wetting, particularly social incontinence. Some of the major reasons for self-wetting are that toilets are far away, roads leading to the toilets are not well maintained, there is a lack of signage and navigation, and there is a lack of safety walking to and at the communal ablution blocks, making children afraid to go to and use them. The toilets used by the children in the study are also not child-friendly. There are not enough facilities and they have not been designed with children in mind. Ullah [30] reported this over a decade ago in older sections of the Rohingya refugee camps.

Studies elsewhere have investigated links between children’s social incontinence and toileting infrastructure. These have indicated that increasing the number of bathrooms, establishing child-friendly toilets, ensuring water availability and improving accessibility on the approach to ablution blocks can encourage children to use facilities, thus reducing social incontinence. For example, a study in the USA found that children who do not have accessible toilets at school are 2.2 times more likely to experience social incontinence than children to have access to accessible toilets [31]. A study in Kenya showed that improvement to school toilet facilities increased the daytime toilet use of children [32].

### Availability of continence management supplies and washing facilities

Our study found that caregivers are busy managing children’s self-wetting and have to fetch water from a distance to wash clothes and bedding, making them feel depressed, irritated, bothered and restless. This is a common experiences of caregivers of people who self-wet in emergency contexts [18].The limited availability of clothes, blankets and diapers made it more challenging for caregivers to manage the leakage issues of children. There is a need for humanitarian actors to more thoughtfully provide non-food items such as mattress protectors, portable toilets for children, and extra soap to the families who have children who self-wet. Médecins Sans Frontières, IRC and IFRC have found the provision of such materials useful to caregivers of children who self-wet in Syria, Iraq, Greece and Honduras [17].

### Knowledge on self-wetting and stigma

The diverse conceptualization and perception of self-wetting and incontinence impact their management in Rohingya camps. Incontinence is considered a disease among the Rohingya community because the leakage of urination and defecation while sleeping is evident even after the consultation with doctors or *Hakeem*. However, prevalence of self-wetting declines with age, reaching a prevalence of 0.5-1.7% by age 16–17 years, with spontaneous cure rates of about 15% yearly between 7 and 12 years and 11% annually between 12 and 17 years [33, 34]. But in emergency settings and LMICs, knowledge about incontinence of both caregivers and health workers is still at an introductory level [3, 17]. We also found that caregivers often do not report self-wetting issues to health and other support workers due to social stigma [18]. Where they are able to hide self-wetting, children often do not inform their caregivers, as has been seen in the Butajira region of southern Ethiopia [9].

To reduce stigma, physical and mental abuse towards children, improve the understanding of continence issues, and prioritise useful interventions by humanitarian actors, there is a need to focus on providing more knowledge to caregivers, children, communities and professionals. A study in Brazil found that caregivers with more education are less likely to severely punish children who self-wet [26]. Humanitarian professionals across the globe have called for further knowledge and training on how to address continence issues in emergency contexts [17].

### Medical intervention

While social incontinence may not need medical intervention (unless there are associated mental health concerns), incontinence that is caused by physical and/or mental health issues may. WASH and protection practitioners are unlikely to be able to directly address the needs of children with incontinence, and should leave this to medical professionals (providing referrals to such services and information where possible). However, providing accessible WASH facilities may assist in removing some of the barriers to children experiencing incontinence, for example, by reducing the time it takes for them to access a suitable toilet.

### Limitations

Our study only included children and their caregivers in two Rohingya camps. However, the camps we chose broadly represented the living situation of most of the Rohingya community, and we also conducted interviews with diverse stakeholders who have a robust understanding of the situation across the Cox’s Bazar camps. Our study did not investigate medical treatment of children with incontinence, and thus did not include health specialists.

## Conclusion

Children living in the Rohingya refugee camps experience self-wetting due to both social and medical incontinence. This study is the first of its kind to speak to both children and their caregivers about this issue, eliciting valuable information on their experiences, as well as suggestions for practical changes. Protection and WASH professionals in emergency settings can better support children and their caregivers through the provision of accessible, close-to-household child-friendly sanitation, providing extra continence management materials such as soap, pads/nappies and mattress protectors, and by both upskilling in their own knowledge around continence issues and communicating this to communities, assisting in better understanding and stigma reduction.

## Data Availability

There are ethical restrictions imposed by the Research Ethics Committee, Faculty of Engineering, University of Leeds, United Kingdom which prevent the public sharing of sensitive minimal data for this study. Data are available upon request from the Research Ethics Committee, Faculty of Engineering, University of Leeds, United Kingdom via email for researchers who meet the criteria for access to confidential data.

## Acknowledgements

The RT would like to acknowledge the many individuals and organisations who contributed to the completion of this project, particularly research trainers and data collectors, administrative staff at all organisations, ethical application reviewers and the project’s Advisory Board. Most importantly they would like to thank the children who participated in the Story Book sessions and their caregivers.

**S1. COREQ (COnsolidated criteria for REporting Qualitative research) Checklist**

**S2. Full codebook**

## References

1. Demaagd, G.A. and T.C. Davenport, Management of urinary incontinence. P t, 2012. 37(6): p. 345–361h.

2. Pizzol, D., et al., Urinary incontinence and quality of life: a systematic review and meta-analysis. Aging clinical and experimental research, 2021. 33: p. 25–35.

3. Rosato-Scott, C., et al., Incontinence: we need to talk about leaks. 2020.

4. Garcia, J.A., J. Crocker, and J.F. Wyman, Breaking the cycle of stigmatization: managing the stigma of incontinence in social interactions. Journal of Wound Ostomy & Continence Nursing, 2005. 32(1): p. 38–52.

5. Gomez Rincon M, Leslie SW, and L. S. Nocturnal Enuresis. StatPearls 2022 [cited 2023; Available from: https://www.ncbi.nlm.nih.gov/books/NBK545181/.

6. Can, G., et al., Child abuse as a result of enuresis. Pediatrics international, 2004. 46(1): p. 64–66.

7. Thom, D.H. and J.S. Brown, Reproductive and hormonal risk factors for urinary incontinence in later life: a review of the clinical and epidemiologic literature. J Am Geriatr Soc, 1998. 46(11): p. 1411–7.

8. Grzeda, M.T., et al., Effects of urinary incontinence on psychosocial outcomes in adolescence. Eur Child Adolesc Psychiatry, 2017. 26(6): p. 649–658.

9. Hafskjold, B., et al., Taking Stock: Incompetent at incontinence—why are we ignoring the needs of incontinence sufferers? Waterlines, 2016: p. 219–227.

10. Mostafaei, H., et al., Prevalence of female urinary incontinence in the developing world: A systematic review and meta-analysis—A Report from the Developing World Committee of the International Continence Society and Iranian Research Center for Evidence Based Medicine. Neurourology and urodynamics, 2020. 39(4): p. 1063–1086.

11. Minassian, V.A., H.P. Drutz, and A. Al-Badr, Urinary incontinence as a worldwide problem. International Journal of Gynecology & Obstetrics, 2003. 82(3): p. 327–338.

12. Walker, G.J. and P. Gunasekera, Pelvic organ prolapse and incontinence in developing countries: review of prevalence and risk factors. International urogynecology journal, 2011. 22: p. 127–135.

13. Paul Abrams, L.C., Adrian Wagg, Alan Wein (Eds,),, Incontinence. 2017, ICI-ICS. International Continence Society: Bristol UK.

14. Wilbur, J., et al., “I’m scared to talk about it”: exploring experiences of incontinence for people with and without disabilities in Vanuatu, using mixed methods. The Lancet Regional Health-Western Pacific, 2021. 14: p. 100237.

15. Rosato-Scott, C.A. and D.J. Barrington, Incontinence in Zambia: initial investigation into the coping strategies of sufferers and carers. Waterlines, 2018. 37(3): p. 190–206.

16. Ansari, Z. and S. White, Managing incontinence in low-and middle income-countries: A qualitative case study from Pakistan. PLOS ONE, 2022. 17(7): p. e0271617.

17. House, S., Chatterton, C., Mapping of support for people living with incontinence in humanitarian contexts, Through the lens of WASH, GBV and ASRH. 2022, Norwegian Church Aid

18. Rosato-Scott, C.A., et al., Urinary incontinence in children aged 5 to 12 in an emergency setting: lessons learned in Ethiopia. Waterlines, 2021. 40(3): p. 179–191.

19. Chess-Williams, R., et al., Chronic psychological stress and lower urinary tract symptoms. LUTS: Lower Urinary Tract Symptoms, 2021. 13(4): p. 414–424.

20. Jurković, M., et al., Refugee status as a possible risk factor for childhood enuresis. International Journal of Environmental Research and Public Health, 2019. 16(7): p. 1293.

21. Rosato-Scott, C., et al., Understanding children’s experiences of self-wetting in humanitarian contexts: An evaluation of the Story Book methodology. PLOS Global Public Health, 2023. 3(5): p. e0001194.

22. Rosato-Scott, C., B.E. Evans, and D.J. Barrington, CHILD-SAN: a new disability-inclusive framework for emergency sanitation for children aged five to 11, based on a systematic review of existing guidance. Journal of International Humanitarian Action, 2021. 6: p. 1–14.

23. Tong, A., P. Sainsbury, and J. Craig, Consolidated criteria for reporting qualitative research (COREQ): a 32-item checklist for interviews and focus groups. International Journal for Quality in Health Care, 2007. 19(6): p. 349–357.

24. UNHCR, Joint Government of Bangladesh - UNHCR Population Factsheet. 2022.

25. Barrington, D., Understanding children and their caregivers’ experiences with incontinence in humanitarian contexts. 2023.

26. Sapi, M.C., et al., Assessment of domestic violence against children and adolescents with enuresis. Jornal de Pediatria, 2009. 85: p. 433–437.

27. Gozmen, S., S. Keskin, and I. Akil, Enuresis nocturna and sleep quality. Pediatric Nephrology, 2008. 23: p. 1293–1296.

28. Rosato-Scott, C., et al., Guidance on supporting people with incontinence in humanitarian and low-and middle-income contexts (LMICs). 2019.

29. Filce, H.G. and L. LaVergne, Absenteeism, educational plans, and anxiety among children with incontinence and their parents. Journal of School Health, 2015. 85(4): p. 241–250.

30. Ullah, A.A., Rohingya Refugees to Bangladesh: Historical Exclusions and Contemporary Marginalization. Journal of Immigrant & Refugee Studies, 2011. 9(2): p. 139–161.

31. Bloom, D.A., et al., Toilet habits and continence in children: an opportunity sampling in search of normal parameters. The Journal of urology, 1993. 149(5): p. 1087–1090.

32. World Bank, Improving health, nutrition and population outcomes in Sub-Saharan Africa: the Role of the World Bank. 2004: The World Bank.

33. Morison, M., H. Staines, and A. Gordon. A Systematic review of the prevalence of Nocturnal Enuresis in children and young people aged 5-18 years and its social impact on the individual and the Family. in Neurourology and Urodynamics. 2004. Wiley-Liss Div John Wiley & Sons inc, 111 river st, Hoboken, NJ 07030 USA.

34. Abrams, P., Cardozo, L, Wagg, A, Wein, A., ed. Incontinence 6th Edition. 2017, International Continence Society.

